# Incidence and risk factors for medical adhesive-related skin injury in catheters of critically ill patients – A prospective cohort study

**DOI:** 10.1101/2022.10.03.22280567

**Authors:** Oleci Pereira Frota, Jéssica do Nascimento Pinho, Marcos Antonio Ferreira-Júnior, Elaine Cristina Fernandes Baez Sarti, Fabiana Martins de Paula, Danielle Neris Ferreira

## Abstract

**Aim:** To investigate the incidence and risk factors for medical adhesive-related skin injury (MARSI) in catheters of critically ill patients.

**Methods:** A prospective cohort study was conducted in adult intensive care units of two Brazilian university hospitals. A total of 150 patients (439 catheters) were included. Skin exposed to the catheter fixation adhesives (central venous, nasogastric, nasoenteral and indwelling urinary) was examined daily by four trained researchers. The patients’ sociodemographic and clinical data were collected from their electronic medical records. The association between independent variables and MARSI was investigated by bivariate statistics, followed by multiple logistic regression.

**Results:** The MARSI incidence was 42% (8.64 MARSIs per 100 patients/day). Advanced age, prolonged hospital stay, dry skin, repetitive adhesive removal, low Braden Scale score and hypoalbuminemia were associated with MARSI (*p* < .05). According to multivariate logistic regression, dry skin increased the chance of MARSI by 5.21 times (odds ratio [OR] 5.21; 95% confidence interval [95% CI] 2.43-11.11), while the Braden Scale score was a protective factor, showing 31% less chance of MARSI for each added score (OR 0.69; 95% CI 0.57-0.85). A higher incidence of MARSI was observed in nasoenteral catheters and in those fixed with adhesive using natural rubber. The MARSI types were predominantly mechanical (70.3%): skin stripping (41.3%), skin tear (26.1%) and tension injury or blister (2.9%).

**Conclusions:** MARSI is a common event in adult intensive care units and most risk factors are modifiable. Preventive actions are potentially capable of reducing incidence, optimizing financial resources and improving clinical results.

## 1. Introduction

Maintaining skin integrity is one of the main indicators of nursing care quality [1] and is closely related to the safety of critically ill patients. A patient admitted to the intensive care unit (ICU) is relatively more susceptible to skin lesions due to greater skin vulnerability and organ dysfunction [2], including medical adhesive-related skin injury (MARSI). MARSI is defined as an “occurrence in which erythema and/or other manifestation of cutaneous abnormality (including, but not limited to, vesicle, bulla, erosion, or tearing) which persists 30 minutes or more after removing the adhesive” [3]. MARSI causes pain, infection, damage to the quality of life of patients, delayed healing and increased length of hospital stay. It therefore causes extra costs to the healthcare system, work overload for the nursing team, emergence of comorbidities and damage to the well-being of patients and families [3]. Although MARSI is preventable, it is relatively common in hospitalized patients, in addition to being underestimated and expensive [4,5].

Several studies on MARSI have been conducted in different settings and populations, such as adult patients admitted to a non-intensive care unit [5], adult patients with acute or chronic wounds in a vascular clinic [6], oncological patients [7], patients requiring peripheral venous insertion catheters [1], surgical patients [8,9] and Neonatal [10], Pediatric [11,12] and cardiac ICU patients [13]. However, a search conducted in PubMed, Web of Science and Scopus found only one study [2] on incidence and risk factors for patients admitted to an adult ICU.

Critically ill patients are very susceptible to MARSI given common conditions such as high exposure to medical adhesives, malnutrition, hemodynamic instability, organ dysfunction, edema, skin abnormalities, and high risk of skin injury by predictive scales [2,14]. In addition, these patients typically need a variety of medical devices for monitoring, diagnosis, and treatment. Urinary, enteral and vascular catheters are among the most used devices which incorporate skin attachment adhesives that are periodically changed according to the adhesive characteristics. Therefore, the use of catheters is a variable which promotes MARSI. However, the density and risk factors for MARSI associated with catheter fixation are currently unknown.

Knowledge of the risk factors associated with MARSI is essential for screening vulnerable people and to establish different preventive care practices. However, there are no validated and widely accepted predictive scales for MARSI, such as the Braden, Norton and Waterlow scales for pressure injuries. Only the instrument by White et al. [15] has been suggested for predicting skin tears. However, it has not been validated, does not appear to be widely used and was not specifically designed for predicting MARSI [16].

Therefore, the aim of this study was to investigate the incidence and risk factors for MARSI in catheters of critically ill patients.

## 2. Methods

### 2.1. Design, setting, period and sampling

This was a prospective cohort, single-group study conducted in two general adult ICUs of two public hospitals (one ICU from each hospital) in Campo Grande, capital of the state of Mato Grosso do Sul, in the central-west region of Brazil from June to August 2019. The hospitals are large, offer healthcare at the tertiary level, perform teaching and research activities, and have a total capacity of approximately 550 beds. The ICU of hospital A has 10 beds, while that of hospital B has 9 beds, totaling 19 beds.

### 2.2. Sampling and selection criteria

The inclusion criteria were: age of at least 18 years old, no MARSI before ICU admission, with catheters fixed to the skin by medical adhesive for at least 24 hours. Patients with congenital dermatological disease or those who requested to withdraw from the study after enrollment were excluded from the study. The analyzed catheters included: central venous catheter (CVC), nasogastric catheter (NGC), nasoenteral catheter (NEC) and indwelling urinary catheters (IUC). Informed consent was obtained from the patient or their legal guardian. The sample was formed by sequential convenience sampling.

### 2.3. Data collection

Data were collected daily by four nurses belonging to the ICUs and who were trained by the main researcher through two theoretical-practical classes on MARSI and the research protocol. Then, nurses were assessed to define and classify MARSI using seven photographs [3] on MARSI types and three [16] on types of skin tears. These lesions were assessed for: (i) identification of MARSI (yes or no); and (ii) classification – Mechanical: skin stripping, tension injury or blister, skin tear (subclassified into type 1, 2 or 3); dermatitis: contact irritation or allergy; other: maceration or folliculitis. The MARSI classification [3] and skin breakdown [16] were defined based on the literature. Strong agreement between data collectors was obtained (Kappa coefficient; K = 0.91).

The following variables were collected from the patients’ medical records: (i) sociodemographic: gender, age, race, height and weight; and (ii) clinical: main diagnosis, comorbidities, length of stay in the ICU, clinical conditions, medications, catheters in use, length of catheter use, Braden Scale score, ICU outcome and results from laboratory tests during follow-up (hemoglobin, hematocrit, albumin and C-reactive protein) performed during the ICU stay, with the means being used for compiling the results. There was no additional follow-up outside the ICU for transferred or discharged patients.

Data were collected on enrolled patients until catheter removal, MARSI identification, ICU discharge, transfer or death. The removal and (re)fixation of the adhesive tapes were performed by the ICU nursing team after the patients’ bath or when necessary, for example in cases of the adhesive being dirty, loss of functionality and removal of the catheter. The skin in contact with the adhesive was examined at the time of adhesive removal and up to 30 minutes after removal. This procedure was observed by the researchers on all days and shifts. Repetitive adhesive removal was defined as an exchange carried out in less than the pre-established time. The adhesives (medical tapes or bandages) were changed when necessary (dirt, detachment, malfunction) or when they reached the maximum permanence time as follows: natural rubber latex based and acrylate every 24 hours; and polyurethane and hydrocolloid every 7 days. The characteristics of these adhesives are described in Table 1.

**Table 1.**
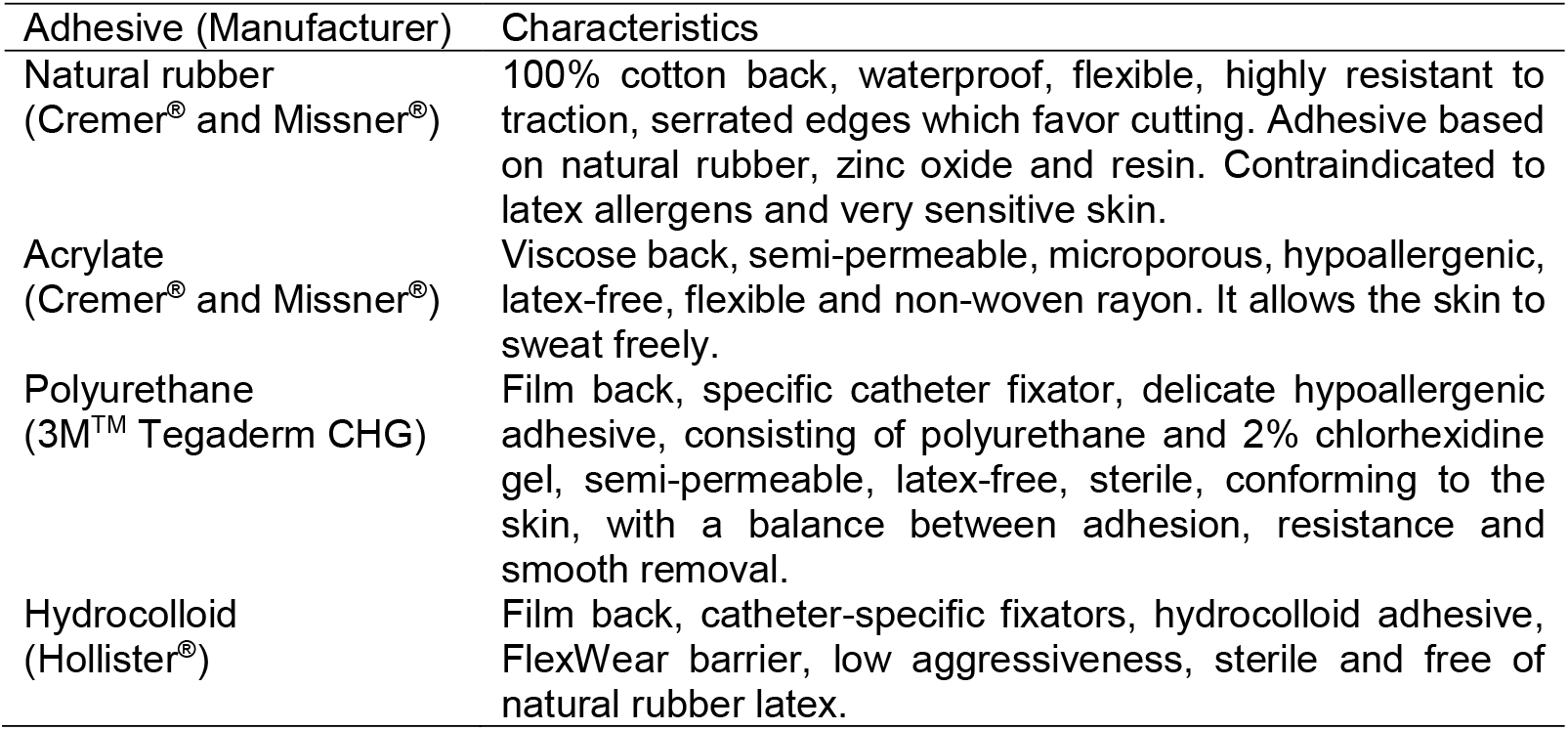
Characteristics of the adhesives.

Considering a three-month surveillance period, the overall care indicators were calculated as follows: (i) MARSI incidence rate: ‘division of the number of MARSI cases’ by ‘number of cases exposed to catheter fixation adhesives’, multiplied by 100; (ii) MARSI incidence rate per 100 catheter days: ‘division of the number of MARSI cases’ by ‘total number of patients who used catheter-day fixation adhesives’, multiplied by 100; and (iii) catheter usage rate per 100 patient-days: ‘division of the number of patients who used a catheter-day’ by the ‘total number of patient-days hospitalized in the period’ multiplied by 100.

### 2.4. Ethical considerations

The study was approved by the Research Ethics Committee of the Universidade Federal de Mato Grosso do Sul, Campo Grande, Mato Grosso do Sul, Brazil (No. 3,096,666) and complied with national and international ethical guidelines for research involving human subjects. Study participants signed the Free Informed Consent Form (FICF) in duplicate, one being delivered to the participant and the other remaining with the researcher.

### 2.5. Statistical analysis

The data were submitted to descriptive and inferential analysis using the Statistical Package for the Social Sciences version 25.0. Non-normal distribution of the data was determined by the Kolmogorov-Smirnov test. The Chi-Squared test for qualitative independent variables and the Student’s t-test for quantitative variables were used to study the association between the independent variables and the MARSI outcome. The significance level was .05. Explanatory variables with a P < .20 value in the bivariate analysis were subjected to multiple logistic regression analysis. The variables were arranged in order of the lowest P-value and analyzed by the Backward Wald Stepwise Method in the modeling process. The model fit was verified by the Hosmer-Lemeshow test. Therefore, the fitted model was adopted as a null hypothesis (H_0_) and the unfitted model as an alternative (H_1_). Thus, model fitting is accepted when P < .05, and the closer to 1.0 the P-value is, the better the fitting quality.

## 3. Results

### 3.1. Sociodemographic and clinical characteristics of patients

A total of 157 patients were admitted to the ICUs of the two hospitals during the study, of which 7 requested to withdraw from the study. Thus, the sample consisted of 150 patients (439 catheters), and their characteristics are described in Table 2.

**Table 2.**
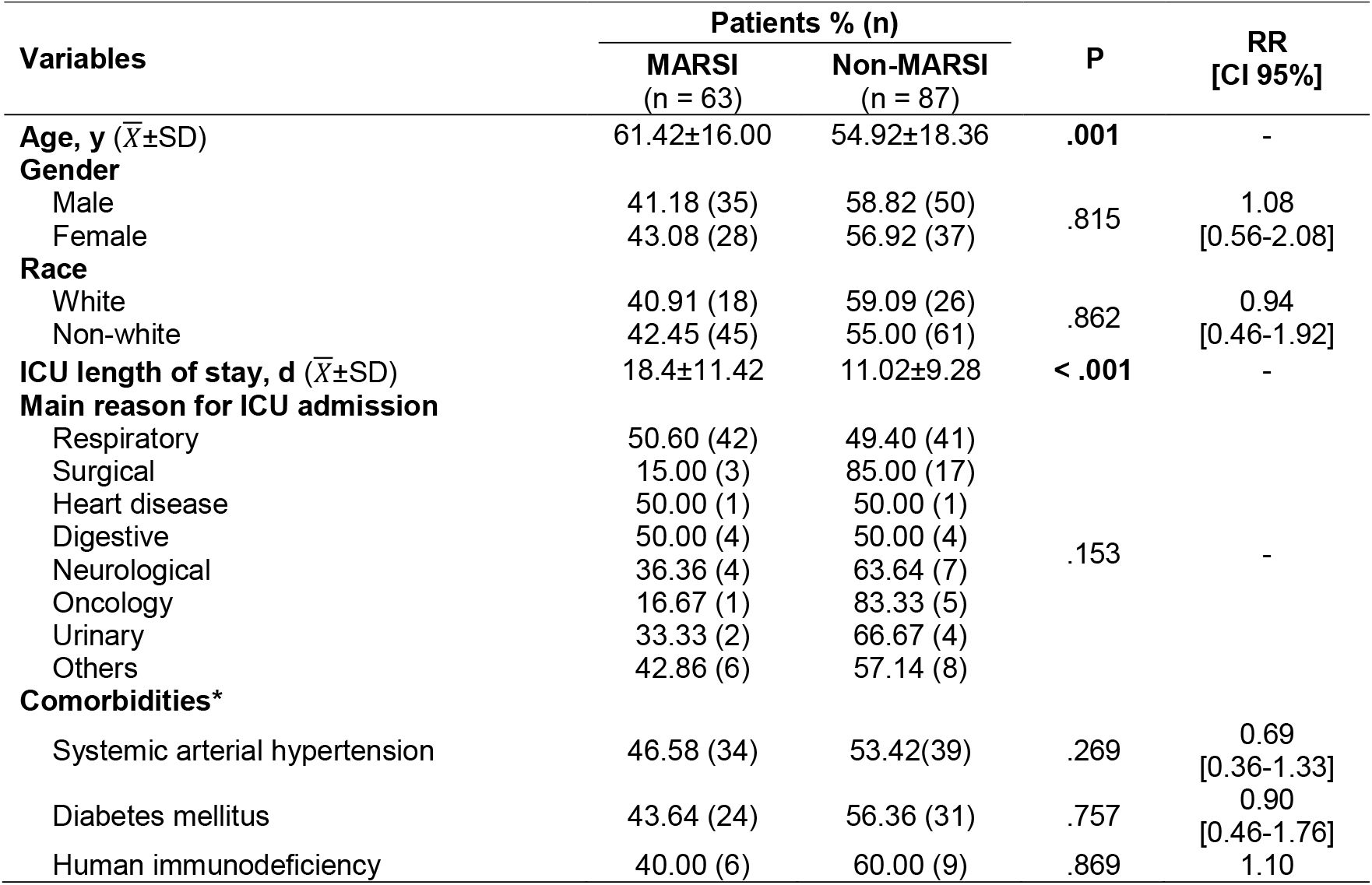
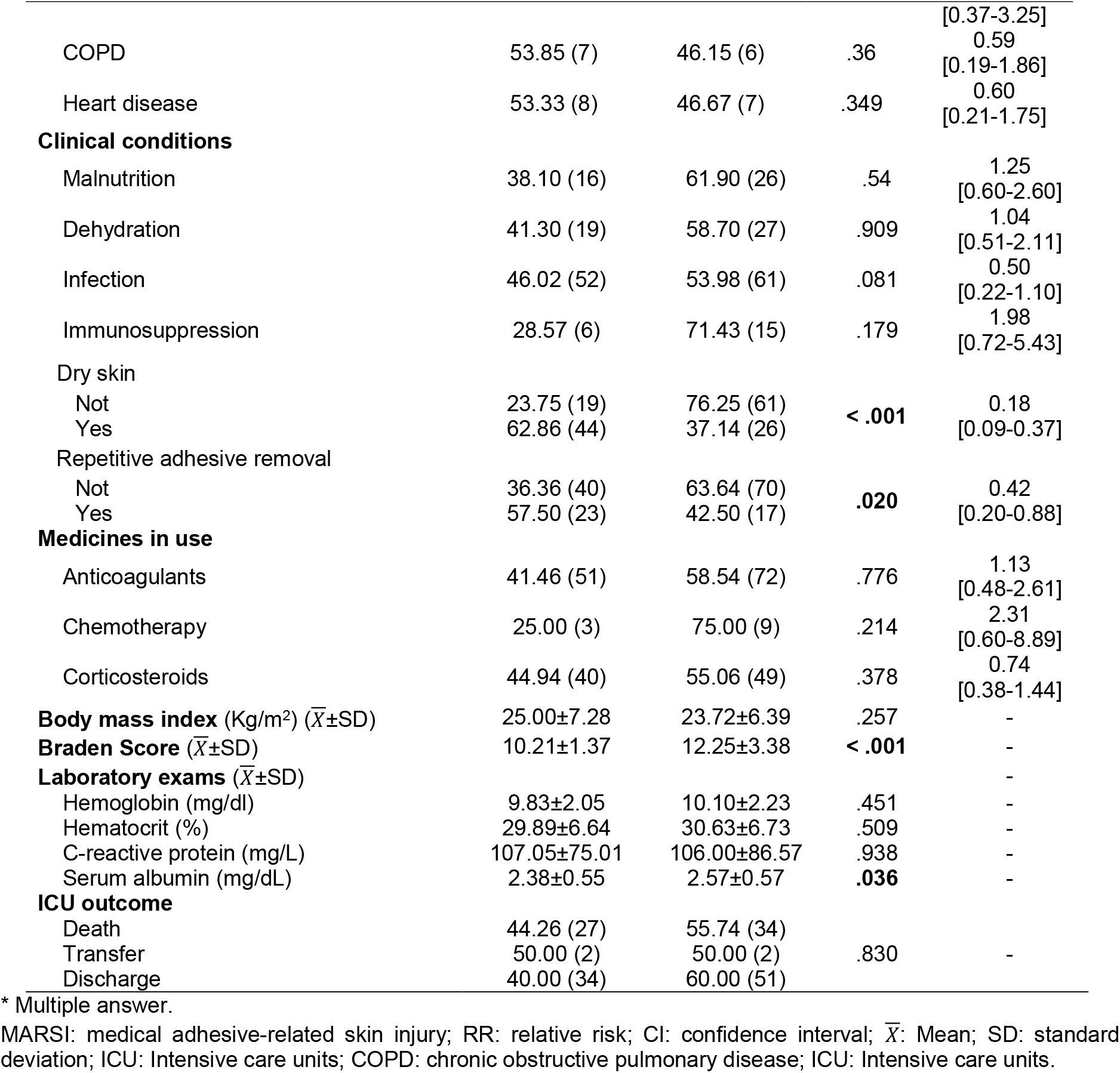
Sociodemographic and clinical variables of patients according to MARSI occurrence.

### 3.2. MARSI incidence

A total of 1596 patient-days were analyzed during the study period, totaling 4,060 catheter-days. The accumulated MARSI incidence rate was 42% ([63 patients who developed MARSI/150 inpatients in the period]), 8.64 per 100 patients/day ([138 MARSI cases/1596 patient-days] x 100), and the cumulative rate of catheter use was 2.54 per 100 patient-days ([4,060 catheter-days in the period/1596 patient-days in the period] x 100) during the three months. The MARSI incidence rate and the catheter usage rate are shown in Figure 1.

**Fig. 1.**
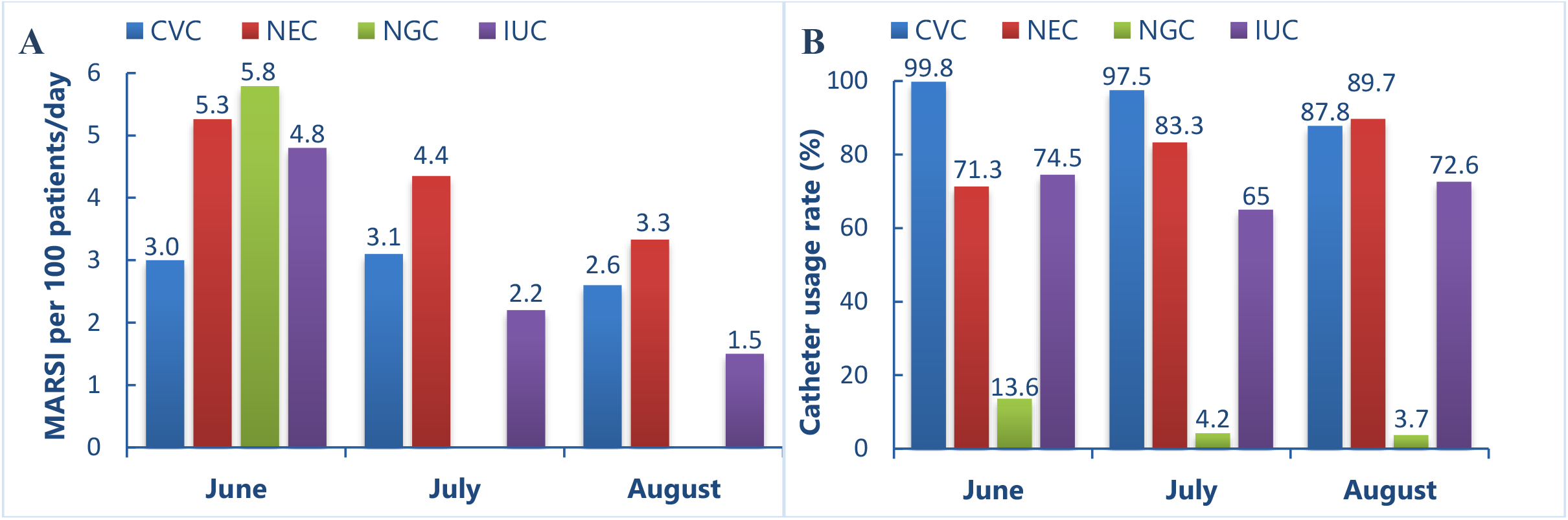
Care indicators for months of research according to type of catheter: **(A)** MARSI incidence rate per 100 patients/day; **(B)** catheter usage rate. MARSI: Medical adhesive-related skin injury; Central venous catheter (CVC); Nasoenteral catheter (NEC); Nasogastric catheter (NGC); Indwelling urinary catheters (IUC).

Both the MARSI incidence and the type of MARSI differed (P < .001) according to the type of adhesive and catheter used. Natural rubber tape was the second most used adhesive in fixing the catheters. However, it was the first cause of injuries (62.32%), being responsible for the highest rate of skin stripping (88.9%) and maceration (91.7%). There was a higher MARSI occurrence related to NEC (39.86%) and CVC (31.88%). NEC stood out for the highest number (P < .001) of skin maceration (80.55%) and skin stripping (61.11%), while skin tearing (52.64%) was more prevalent in the use of CVC (P < .001). There were no cases of allergic dermatitis or folliculitis, and 93.5% of the MARSI burden was skin tearing (41.3%), skin stripping (26.1%) and maceration (26.1%) (Table 3).

**Table 3.** MARSI incidence and type according to the adhesive tape and catheter type (n=439).

| MARSI<br>occurrence<br>and type (n) | Type of adhesive or catheter |  |  |  |  |  |  |  |  | P* |
| --- | --- | --- | --- | --- | --- | --- | --- | --- | --- | --- |
|  | Adhesive n(%) |  |  |  | P* | Catheter n(%) |  |  |  |  |
|  | Acrylate | Natural rubber | Hydrocolloid | Polyurethane |  | CVC | IUC | NEC | NGC |  |
| MARSI (n=439) |  |  |  |  |  |  |  |  |  |  |
| No (301) | 134(44.51) | 58(19.27) | 61(20.27) | 48(15.95) | < .001 | 107(35.55) | 109(36.21) | 67(22.26) | 18(5.98) | .001 |
| Yes (138) | 47(34.06) | 86(62.32) | 3(2.17) | 2(1.45) |  | 44(31.88) | 25(25.36) | 55(39.86) | 4(2.90) |  |
| MARSI type (n=138) |  |  |  |  |  |  |  |  |  |  |
| M1 (36) | 4(11.11) | 32(88.89) | - | - | < .001 | 6(16.67) | 7(19.44) | 22(61.11) | 1(2.78) | < .001 |
| M2 (4) | 4(100.00) | - | - | - | .007 | 3(75.00) | 1(25.00) | - | - | .109 |
| M3 (57) | 34(59.67) | 20 (35,08) | 2(3,50) | 1(1,75) | < .001 | 30(52,64) | 23(40,35) | 4(7,01) | - | < .001 |
| M3-I (4) | 3(75.00) | 1(25.00) | - | - |  | 2(50.00) | 2(50.00) | - | - |  |
| M3-II (10) | 6(60.00) | 3(30.00) | - | 1(10.00) |  | 7(70.00) | 2(20.00) | 1(10.00) | - |  |
| M3-III (43) | 25(58.14) | 16(37.21) | 2(4.65) | - |  | 21(48.83) | 19(44.19) | 3(6.98) | - |  |
| D1 (5) | 2(40.00) | 1(20.00) | 1(20.00) | 1(20.00) | .895 | 3(60.00) | 2(40.00) | - | - | .141 |
| D2 (0) | - | - | - | - | 1.00 | - | - | - | - | 1.00 |
| O1 (36) | 3(8.33) | 33(91.67) | - | - | < .001 | 2(5.56) | 2(5.56) | 29(80.55) | 3(8.33) | < .001 |
| O2 (0) | - | - | - | - | 1.00 | - | - | - | - | 1.00 |

\* $\chi^2$ test or Fisher's exact test.
MARSI: medical adhesive-related skin injury; CVC: central venous catheter; IUC: indwelling urinary catheters; NEC: nasoenteral catheter; NGC: nasogastric catheter.
**Note:** M: mechanical (M1: skin stripping; M2: tension injury or blister; M3: skin tear [M3-I: skin tear type 1; M3-II: skin tear type 2; M3-III: skin tear type 3]); D: dermatitis (D1: irritant contact dermatitis; D2: allergic dermatitis); O: other (O1: maceration; O2: folliculitis).

### 3.3. Associated risk factors

The explanatory variables with P-value < .20 in the bivariate analysis – age, ICU length of stay, main reason for ICU admission, infection, immunosuppression, dry skin, repetitive adhesive removal, Braden Score and serum albumin – (Table 2) were selected for multiple logistic regression by the Backward Wald Stepwise method. The final model included two variables: dry skin and Braden Scale score, with the chance of patients with dry skin to develop MARSI being 5.21 times greater than those without dry skin (odds ratio [OR], 5.21; 95% confidence interval [CI], 2.438-11.113; P < .001), and there is a 31% decrease in the patient’s chance of presenting MARSI with each score added to the Braden Scale (OR, 0.695; 95% CI, 0.568-0.851; P < .001). From the scores in the table, it was found that the model was adequate (Table 4).

**Table 4.** Multivariate logistic regression analysis of risk factors for medical adhesive-related skin injury incidence (Stepwise method) (n=439).

| Variable | $\beta$ | SE | Wald | P | OR (95% CI) |
| --- | --- | --- | --- | --- | --- |
| Dry skin | 1.650 | 0.387 | 18.176 | **< .001** | 5.21(2.438-11.113) |
| Braden Scale | -0.364 | 0.103 | 12.454 | **< .001** | 0.695 (0.568-0.851) |
| Constant | 2.824 | 1.095 | 6.649 | **0.010** | - |

$\beta$ : Beta Coefficient; SE: Standard Error; Wald: Logistic regression test; OR: Odds ratio; CI: Confidence interval.
**Note:** Waste $\chi^2$ (Overall Statistics): $P = .524$ ; Hosmer-Lemeshow: $P = .142$ ; Hit percentage: 73.33%; Cox and Snell: .244; Nagelkerke: .328

## 4. Discussion

### 4.1. Incidence

The MARSI cumulative incidence rate in this study (42%) was higher than that found in non-intensive inpatient units for adults: 15.5% in long-term care units [17] and 5.8% in outpatient vascular clinics [6]. MARSI prevalence studies with adults also report lower frequencies outside the ICU [1,5,7] and cardiac ICU [13]. This finding can be explained by the fact that most critical patients present the nursing diagnosis of risk of impaired skin integrity [18] as secondary to organic disorders, subsequent treatment (inotropes, vasopressors) and management (immobility, bed bound). Additionally, practically three-quarters (74.03%) of the catheters were fixed with relatively more aggressive adhesives to the skin: acrylate (41.23%) and natural rubber (32.80%). In addition to the chemical properties and mechanisms of adhesion to the skin [19,20], such adhesives probably showed greater association with MARSI because they were removed and refixed a greater number of times: every 2 days, versus 7 days for the hydrocolloid and polyurethane.

A prospective cohort study [2] performed with 356 adults admitted to two surgical ICUs (general and vascular) in Beijing, China, found a MARSI incidence of 10.96%. This is lower than that found in our study, which can be justified by the profile of the patients. Our study was conducted in general ICUs, and only 13.3% of patients were hospitalized for surgical reasons. Surgical ICUs usually admit less severe patients than clinical ICUs, therefore they often have fewer organ dysfunctions which increase the patient’s susceptibility to MARSI, namely: fever, sepsis, circulatory shock, respiratory failure, etc.

### 4.2. Associated risk factors

Advanced age was associated with an increased risk of MARSI. The water content and skin elasticity are reduced [2] with advancing age, and the dermoepidermal junction becomes flattened [21]. As a result, aged skin is thinner, parched, and fragile [22], and therefore more vulnerable to MARSI.

Studies with adult [2] and pediatric [11] patients report that longer ICU stay is a risk factor independent of MARSI, which is consistent with this study. This can be explained by the greater exposure to the successive removal of adhesives and the incidence of risk factors such as: malnutrition, infections, and certain medications (i.e. anti-inflammatory, anticoagulants, long-term corticosteroid use) [3], among others. Thus, in addition to the application of preventive protocols, a goal of the health service should be timely removal of devices fixed to the skin by adhesives when there is no longer a clinical need for the device.

Patients who underwent repetitive adhesive removal had more MARSI than those who did not. This can be explained by the fact that repetitive removal may peel off the superficial epidermal layers of the skin and/or trigger an inflammatory or allergic reaction [23]. In addition to being attributed to repeated dressing applications, the trauma is generally associated with inappropriate use of dressings with very aggressive adhesives [24]. Therefore, in high-risk patients such as those in ICU, the use of a soft removal adhesive that allows replacement over long intervals is recommended. Silicone adhesives – the newest available class – meet these criteria, as they are characterized by high and constant adhesion (without adhesion increasing over time), smooth removal [19] and low stratum corneum removal, maceration and pain on removal, latex-free and hypoallergenic [3,25]. Further investigations may determine the effect of using less aggressive adhesives in ICUs.

Hypoalbuminemia can cause homeostatic imbalances, edema, anasarca, and as a consequence can favor skin lesions, as has been well documented for pressure injuries [26]. An important consensus in MARSI recommends using soft, elastic adhesive products if swelling is anticipated at the device fixation site [3]. Similar to this study, Zhang et al. [2] demonstrates that hypoalbuminemia is a risk factor for MARSI. It is recommended that this parameter be considered to identify patients at high risk for MARSI in ICUs and to institute a differentiated prevention protocol.

Dry skin at the contact site with the adhesive was an independent risk factor, as it increased the chance of MARSI by 5.21 times. It is the most common clinical manifestation among dermatological diseases [22], a risk factor for skin stripping [3] and frequently present in older adults [27]. However, it is a modifiable factor through preventive actions such as periodically moisturizing the skin, combatting excessive moisture, applying essential fatty acids, avoiding the use of adhesives [3], using adhesive removers, skin protector and avoiding the use of alcohol-based products, perfumes or aggressive soaps [25].

Corroborating with the literature [3,11], the Braden Scale score was an independent protection factor for MARSI, meaning that the higher the score, the lower the probability of MARSI. The patient’s chance of developing MARSI in pediatric ICUs [11] decreased by 7% for each point added to the Braden Q score against 24% in adults hospitalized in Chinese ICUs [3], and 31% in this study. Although designed to predict the risk of pressure injury, these studies support the “off-label” use of the Braden Scale to identify patients at high risk for MARSI and recommend early interventions. However, it is prudent to build a prediction scale specific to MARSI.

### 4.3. Strengths and implications

Since researchers are in the early stages of reporting and understanding MARSI and its causes [4], this investigation provides an opportunity to understand MARSI as a common ICU problem and offers subsidies for identifying vulnerable people and to develop predictive scales and care protocols. This study found that: advanced age, prolonged hospital stay, dry skin, repetitive adhesive removal, low Braden Scale score and hypoalbuminemia contribute to the development of MARSI; (ii) catheters fixed with adhesive based on natural rubber, zinc oxide and resin are more susceptible to MARSI and (iii) nasoenteral and central venous catheters are the main catheters associated with MARSI.

### 4.4. Limitations

This study was conducted in only two adult ICUs, included only four types of adhesives, and the number of exposures to the adhesives and the severity of patients and injuries were not collected for analysis. Further studies, especially multicenter ones, are needed to determine the overall MARSI incidence in critically ill patients, as well as risk factors. They should consider (or standardize) the following influences: the contact area size of the adhesives with the skin; and the procedures for fixing and removing the adhesive tapes.

## 5. Conclusion

MARSI is a common and harmful event for the safety of critically ill patients, but it is potentially preventable. Prevention is the main nursing action to face this problem and depends on the professionals’ awareness and screening of vulnerable people. This study contributes to this, as it identified the incidence and risk factors such as advanced age, long hospital stay, hypoalbuminemia, dry skin and those at risk according to the Braden Scale score. Our results provide an empirical basis for care and administrative guidelines, as well as for future epidemiological and clinical research in ICUs.

## Data Availability

All data produced in the present study are available upon reasonable request to the authors

